# Association between RT-PCR Ct Values and COVID-19 New Daily Cases: A Multicenter Cross-Sectional Study

**DOI:** 10.1101/2020.12.07.20245233

**Authors:** Abdulkarim Abdulrahman, Saad I. Mallah, Abdulla Alawadhi, Simone Perna, Essam M. Janahi, Manaf M AlQahtani

**Author notes:** **Correspondence: *Lt. Col. Dr. Manaf Al-Qahtani***, MB Bch BAO(Ireland), MMM(USA), FACP(USA), FRCPC(Canada), Head of Infection Control Unit and Microbiology Department, Bahrain Defence Force Hospital, Chairperson of the COVID-19 Control Room, Member of the National Taskforce for Combating the Coronavirus (COVID-19), Associate Professor of Medicine, Royal College of Surgeons in Ireland – Bahrain. **Conflict of interest Disclosures:** The authors declare no conflict of interest.

## Abstract

**Introduction:** Proactive prediction of the epidemiologic dynamics of viral diseases and outbreaks of the likes of COVID-19 has remained a difficult pursuit for scientists, public health researchers, and policymakers. It is unclear whether RT-PCR Cycle Threshold (Ct) values of COVID-19—or any other virus—as indicator of viral load, could represent a possible predictor for underlying epidemiologic changes on a population level.

**Objectives:** To investigate whether population-wide changes in SARS-CoV-2 RT-PCR Ct values over time are associated with the daily fraction of positive COVID-19 tests. In addition, this study analyses the factors that could influence the RT-PCR Ct values.

**Method:** A retrospective cross-sectional study was conducted on 63,879 patients from May 4, 2020 to September 30, 2020, in all COVID-19 facilities in the Kingdom of Bahrain. Data collected included number of tests and newly diagnosed cases, as well as Ct values, age, gender nationality, and symptomatic status.

**Results:** Ct values were found to be negatively and very weakly correlated with the fraction of daily positive cases in the population r = −0.06 (CI95%: −0.06; −0.05; p=0.001). The R-squared for the regression model (adjusting for age and number of daily tests) showed an accuracy of 45.3%. Ct Values showed an association with nationality (p=0.012). After the stratification, the association between Ct values and the fraction of daily positive cases was only maintained for the female gender and Bahraini-nationality. Symptomatic presentation was significantly associated with lower Ct values (higher viral loads). Ct values do not show any correlation with age (p=0.333) or gender (p=0.522).

**Conclusion:** We report one of the first and largest studies to investigate the epidemiologic associations of Ct values with COVID-19. Ct values offer a potentially simple and widely accessible tool to predict and model epidemiologic dynamics on a population level. More population studies and predictive models from global cohorts are necessary.

## Introduction

Coronavirus Disease 2019 (COVID-19) has ravaged the world by surprise over a period of half a year. According to the latest World Health Organization (WHO) report on 7 November 2020, there were 48,534,508 confirmed cases and 1,231,017 deaths worldwide, with 82786 confirmed cases and 328 deaths reported in Bahrain. When discussing the epidemiology of a virus, a thorough understanding of its transmission is necessary. SARS-CoV-2, the causative agent of Covid19, primary mode of transmission is through respiratory droplets and direct or indirect contact routes^1^. This occurs when a person is in close contact with infected person who has respiratory symptoms or exchanges in droplet-expelling processes such as talking or singing. The non-infected individual thus exposes their mucosa and conjunctiva to SARS-CoV-2 infected droplets. Indirect contact through fomites may also be a source of infection when transferred from surfaces that have been contaminated with infected droplets, to the mouth, nose, or eyes. Although the epidemiology of SARS-CoV-2 doesn’t support airborne transmission in public settings, an increasing number of publications are pointing to such route. Other less common routes for infection may include different bodily fluids (e.g. sexual, faeces, and blood). The exact role of asymptomatic and pre-symptomatic transmission in the COVID-19 outbreak is not fully comprehended as of yet, but is believed to have an important contribution to the viruses’ rapid spread. SARS-CoV-2 RNA may be detected 1-3 days prior to symptom onset. The viral load is highest around the day of symptom onset, followed by a gradual decline over time. RT-PCR positivity may last for 1-2 weeks in the case of asymptomatic patients, and up to 3 weeks for those with mild to moderate disease^2^.

It is important to differentiate between viral load and infectious dose, and the inter-related implications of both. The infectious dose of a virus is the dose at which if a person is exposed to it would be sufficient to cause an infection. On the other hand, viral load is the amount of virus particles that the infected patient carries in their system. The higher the viral load of a patient, the more virus is shed, which therefore leads to a higher chance of others being exposed to the infectious dose needed to acquire the disease. As with viruses in general, the viral load of COVID-19 patient can be deduced from the Cycle threshold (Ct) value of the RT-PCR test conducted on the obtained sample. The RT-PCR test amplifies the viral RNA from the patient’s sample until it is at a detectable concentration that exceeds the threshold value. The number of cycles necessary for that to take place is known as the Ct value. Thus, the lower the Ct value of a patient’s sample, the higher the viral load, whereas the higher it is the lower the viral load^3^. According to one study, patients with severe symptoms presented with 60 times higher viral load and prolonged viral shedding than patients with mild symptoms, which may indicate a role for viral load not only in diagnosis, but rather prognosis and transmission/contagiousness. Studies showed that lower Ct values were associated with a higher probability of a positive viral culture. In one study, infectivity (defined as growth in cell culture) was significantly reduced when RT-PCR Ct values were over 24^4^. On the other hand, La Scola et al. (2020) have reported that patients with Ct values ≥34 are non-infectious and can be discharged from hospitals or care centers^5^. The implications of Ct values or viral load, if any, on the collective population from an epidemiologic perspective has not been adequately studied yet.

Proactive forecasting of the disease’s epidemiology has remained a distant desire and puzzling enigma for scientists, public health researchers, and policymakers. Unlike previous outbreaks, the impact of the COVID-19 pandemic has transcended the medical and clinical setting, bringing forth major global disruption to public life in the social and professional settings, as well as serious economic repercussions on national and global levels^6^. Countries’ deployment of their health resources and response preparedness has also been a source of concern, due to the unexpected nature and trajectorial development of the pandemic^7^. For these reasons, the prediction or forecasting of an upcoming surge in COVID-19 cases is an important item on the agenda of the scientific community and decision-makers. The daily number of positive cases on a global level has been widely variable. For instance, September 1st, 2020, has witnessed the highest number of positive COVID-19 cases since the beginning of the outbreak with 439,070 cases. The day previous, however, recorded 264,107 cases, while the day after recorded only 170,846; the lowest since the 30th of June. Forecasting attempts have been utilized to a fair degree of accuracy so far. The US Centers for Disease Control and Prevention (CDC) publishes multiple forecasts modelled by independent institutions, predicting the daily number of new cases in the USA. The models use a variety of data types besides those directly related to COVID-19, including demographic and mobility data, as well as different methods and estimates of the impact of public health interventions/policies and how these levels of precautions will change^8^. The previous forecasts when compared to actual data do fairly well in terms of general trend and values to be expected^9^. However; the multivariable models are fairly complex and may not be readily available for use in other countries.

Therefore, this study objective is to investigate whether population-wide changes in SARS-CoV-2 RT-PCR Ct values over time, are correlated with the number of new COVID-19 cases in the future. Our hypothesis is based on that higher viral load indicates increased viral shedding, increased risk of exposure to an infectious dose, and thus a higher degree of transmissibility. Hence, a decrease in Ct value could predict an increase or a surge in the number of positive cases. In order to develop a predictive model, other epidemiological factors associated with Ct values were also investigated. The ability to predict population changes in the number of COVID-19 positive cases ahead of time using a widely available and simple metric can be very helpful. Public health measures and policies can be proactively tailored to control and manage the outbreak in an informed and prophylactic manner.

## Methods

### Study Population & Data Collection

A retrospective study was conducted on 63,879 patients from May 4, 2020 to September 30, 2020, in all COVID-19 facilities in the Kingdom of Bahrain. Data on the number of tests and newly diagnosed cases were obtained from the National COVID-19 Task force’s centralized database. Data collected included age, sex, nationality, symptomatic status. Ct Values from newly diagnosed cases in this time period were obtained from the corresponding medical records.

### Diagnosis and SARS-Cov-2 RNA Amplification

All cases were diagnosed as COVID 19 based on RT-PCR tests of nasopharyngeal samples. The nasopharyngeal samples were transferred to a viral transport media immediately after collection and transported to a COVID-19 laboratory for testing. Diagnosed cases were a heterogeneous sample of tested patients, including symptomatic individuals, close contacts, travelers, healthcare worker screening, and random population testing. The RT-PCR tests were conducted using different kits : Thermo Fisher Scientific (Waltham, MA) TaqPath 1-Step RT-qPCR Master Mix, CG (catalog number A15299), SuperScript™ III Platinum™ One-Step qRT-PCR Kit (catalog number 11732020), LightMix1 Modular SARS-CoV (COVID19) (TIB MOLBIOL, Berlin, Germany). The RT-PCR was conducted on the Applied Biosystems (Foster City, CA) 7500 Fast Dx RealTime PCR Instrument, LightCycler® 480 Instrument II (Roche Molecular Systems, Inc). The assay used targeted the E gene. If positive, the sample was confirmed by RdRP and N genes. The E gene Ct value was reported and used in this study. Ct Values >40 were considered negative.

### RT-qPCR Analytical Validation

Positive and negative controls were included for quality control purposes. Linear range was determined using serial log_10_ dilutions of standard RNA and was established between 3·5 × 10^3^ to 3·5 × 10^12^ copies of RNA/mL, with amplification efficiency of 87% and with a regression R^2^ value higher than 0.99.

### 3. Data Handling and Statistical Analysis

Statistical analyses were performed using SPSS 25.0 (SPSS, Inc., Chicago, IL, USA). The sample size was not calculated as this was a pilot study. Mean and standard deviation (SD) for all variables were given as descriptive statistics. Continuous variables were calculated between groups by means and SDs and frequencies. Categorical variables were reported as frequencies and percentages. The data was grouped into a daily set consisting of 150 days, and a weekly set, consisting of 21 equal weeks starting on May 4^th^ and ending on September 27^th^.

According to the Kolmogorov-Smirnov normality test, all variables were normally distributed. The main endpoint was assessed with the Pearson’s correlation analysis, subsequently stratified for gender and nationality. Linear regression analysis was applied in order to identify the best prediction model for identify the new daily cases, including as regressors, Ct values, new daily tests and age.

## Ethics Approval

The study was approved by the National COVID-19 Research and ethics committee, code number: CRT-COVID2020-082.

## Results

### Characteristics of the Study Population

The total number of positive cases identified between May 4, 2020 and September 30, 2020 (5-month period) was 63,883. Mean age of the individuals was 34.02 years (±16.403), with a male-majority (67.24%, n=42953). Bahrainis represented majority of the cohort (53.68%). Only 35.25% (n = 22,519) of the cohort was symptomatic. The mean number of tests conducted per day was 9140 tests (± 1860), while the mean number of newly diagnosed cases per day was 493 (± 142). Mean proportion of positive tests was therefore 5.43% (± 1.29). The mean Ct values of the population is 25.49 (95%CI: 25.45 - 25.52). Minimum recorded Ct value was 10.16, while the maximum was 40.38. The weighted mean Ct value in the 25^th^ percentile was 22, and 28.89 in the 75^th^ percentile. Table 1 summarizes the sample characteristics.

**Table 1.** General characteristics of the sample. This study enrolled a total sample of 63,883 patients. Bahraini (53.7%), Expats (46.3%) with mean of age of 33.97±16.60 years old. In total, 32.8% were females while 67.2% were males (67.2%).

| Variable | Frequency | Mean ; SD |
| --- | --- | --- |
| Total sample | 63883 |  |
| Age | | $33.97 \pm 16.60$ |
| Gender | Female (32.8%) (20930)<br>Male (67.2%) (42953) |  |
| Nationality | Bahraini (53.7%)<br>Expats (46.3%) |  |
| Symptoms | No 64% (41364)<br>Yes 36% (22519) |  |
| CT Values | | $25.46 \pm 4.69$ |
| Number of daily tests | | $9258 \pm 1866$ |
| Total cases | | $498 \pm 144$ |

### Primary Endpoint: Relationship between Ct Value and New Dailey Cases

Analysis of the association between Ct values and ratio of new daily cases to daily tests (percentage of positive cases), reveals a significant correlation coefficient of -0.06 (95%CI: -0.06 to -0.05, p<0.001) (Figure 1).

**Figure 1.**
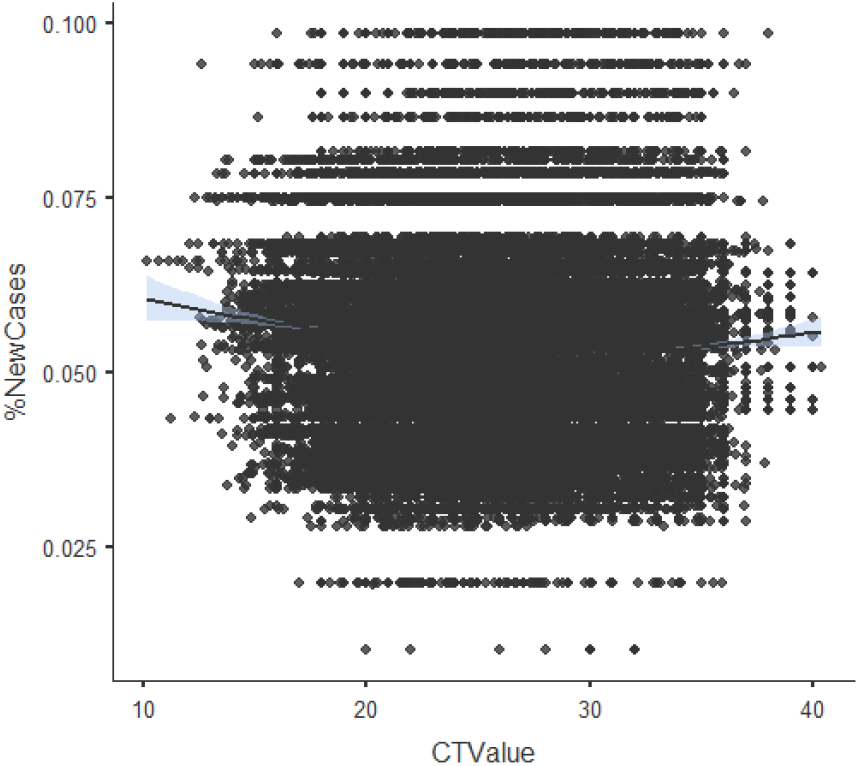
Figure 1 shows the Pearson’s correlation between Ct values and Ratio new daily cases/new tests. The correlation coefficient was r=−0.06 (CI95%: −0.06;−0.05; p=0.001)

### Factors Affecting Ct Value

Analysis of the association between Ct values and patients’ age returned a non-significant Pearson correlation (r = 0.00, p = 0.455) (Figure 2). Nationality was found to be significantly associated with Ct value as illustrated in Figure 3. Bahraini nationals had significantly higher mean Ct values than their non-Bahraini/expat counterparts (25.53 vs 25.44, p = 0.014). As for gender, no association was found with Ct values (Figure 4). However, males did report a lower Ct value on average compared to females (25.48 vs 25.510, p = 0.522). Finally, symptomatic status was found to be significantly associated with lower Ct values (p = 0.001), when compared to asymptomatic status (24.93 vs 25.79) (Figure 5).

**Figure 2.**
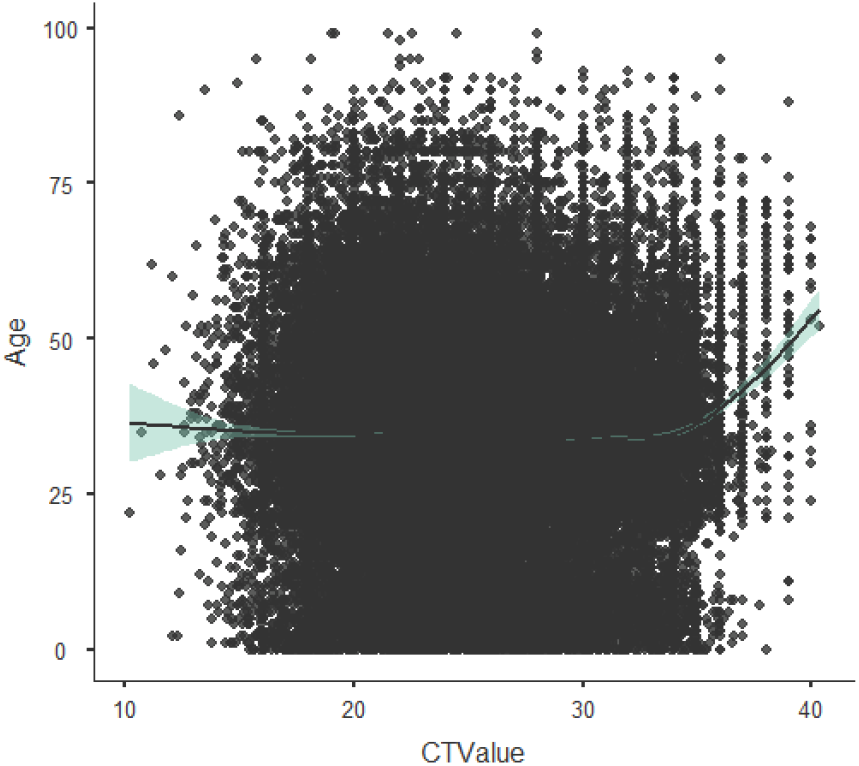
Figure 2 shows the effect of age on the Ct values. Ct values are not affected by age: the r is equal to 0.000 and the p value is not statistically significant (p=0.455).

**Figure 3.**
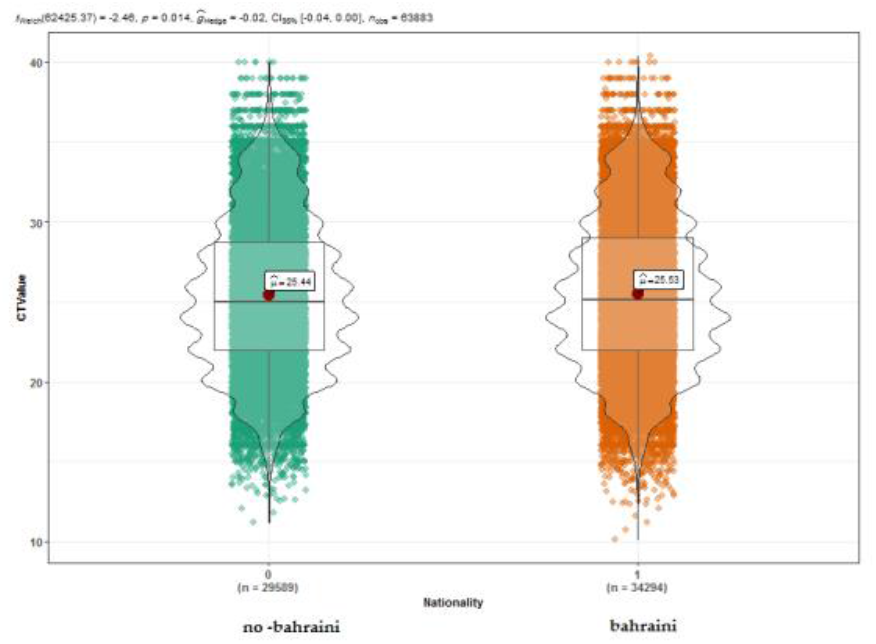
Effect of nationality on the Ct values. Ct values are affected by nationality (p=0.014). Bahraini nationality have a higher Ct values than non-Bahraini (25.53 versus 25.44) In figure 4 Is reported the effect of gender on the Ct values. Ct values are not affected by gender (p=0.522). Males have lower Ct values compared to females (25.48 versus 25.51).

**Figure 4.**
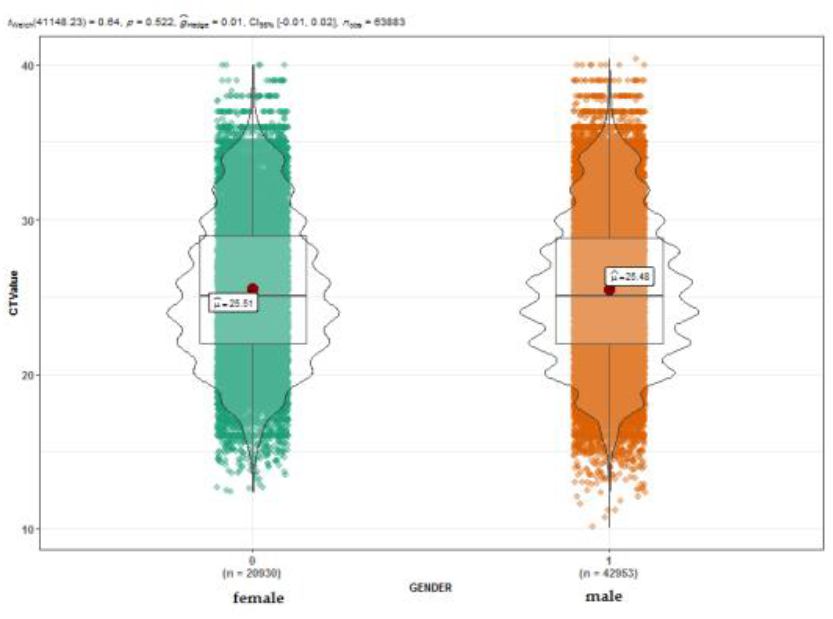
Effect of nationality on the Ct values. Ct values are affected by nationality (p=0.014). Bahraini nationality have a higher Ct values than non-Bahraini (25.53 versus 25.44) In figure 4 Is reported the effect of gender on the Ct values. Ct values are not affected by gender (p=0.522). Males have lower Ct values compared to females (25.48 versus 25.51).

**Figure 5.**
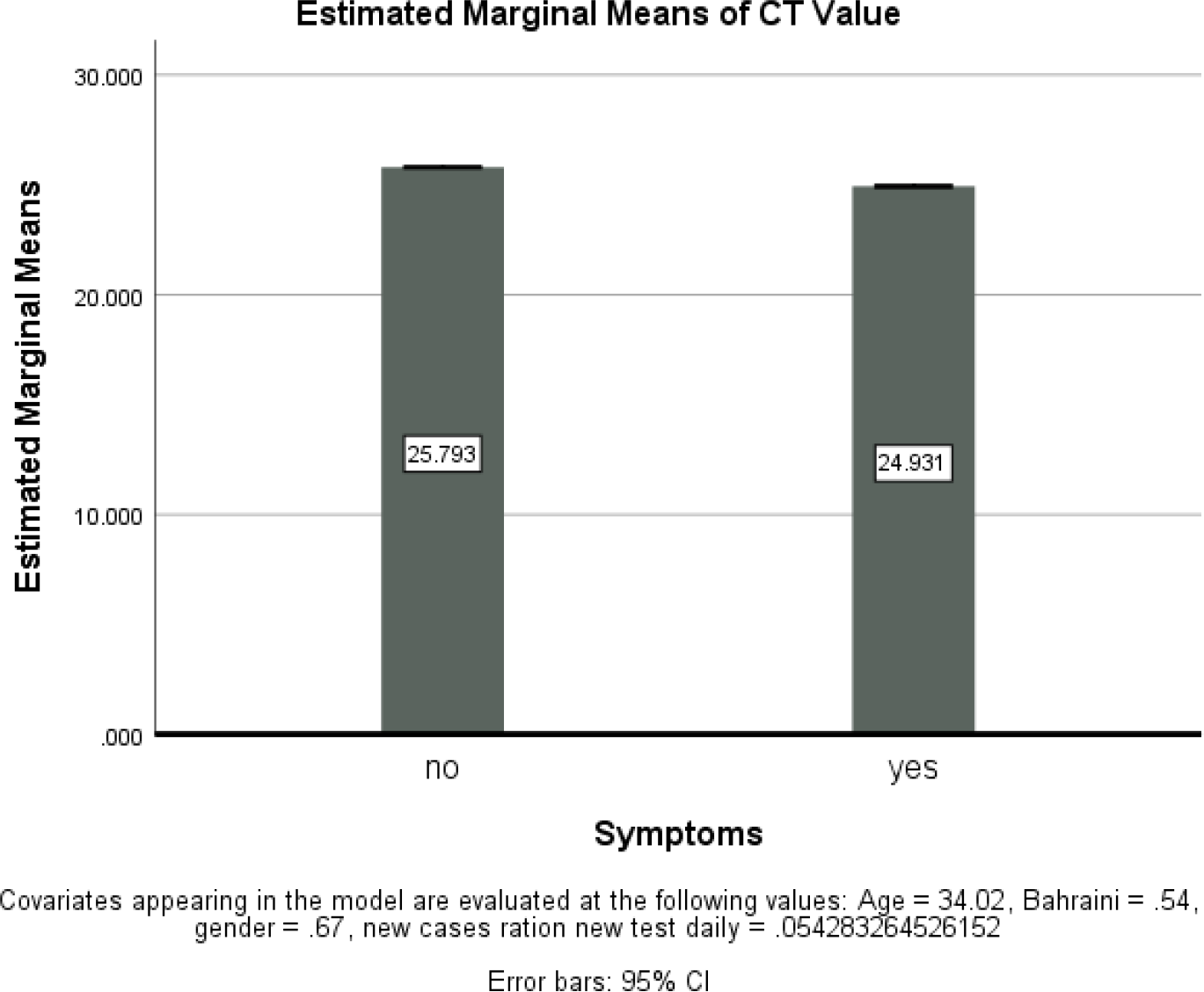
The mean Ct values of Symptomatic versus Asymptomatic Patients. Between the 2 groups there is a statistically significant difference (p=0.001). Symptomatic patients show lower level of Ct values (Symptomatic=24.931 versus Asymptomatic=25.793).

### Predictive Model Parameter Identification

A regression model on association between Ct values (predictor) and new daily cases (outcome) was also conducted (Table 2, Figure 6). The covariates included age and daily tests. Pearson’s correlation returned a strong r value of 0.673, and an r^2^ of 0.453. The model’s level of prediction was 45.3%. Table 2 shows the regression model, where Ct value had a significant negative correlation with new daily cases (coef = -1.82, p<0.001). Daily tests was positively correlated with daily new cases (coef = 0.05, p<0.001). Age was not found to be associated with new daily cases.

**Figure 6.**
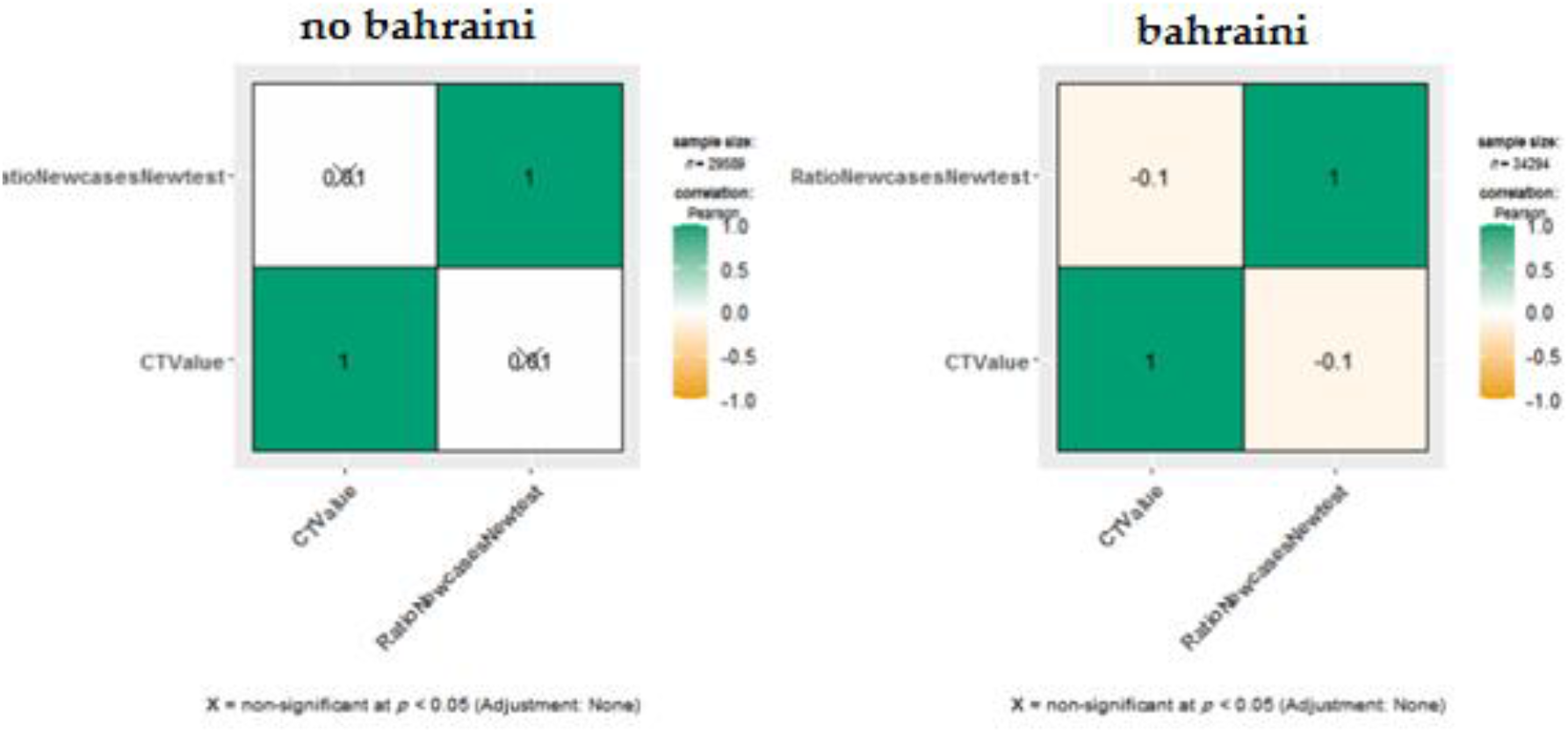
Figure 6 reports the effect of nationality on the relationship between Ct values and ratio of new cases/new tests. A negative correlation was found between Bahraini cases (p = 0.001) only.

**Figure 7.**
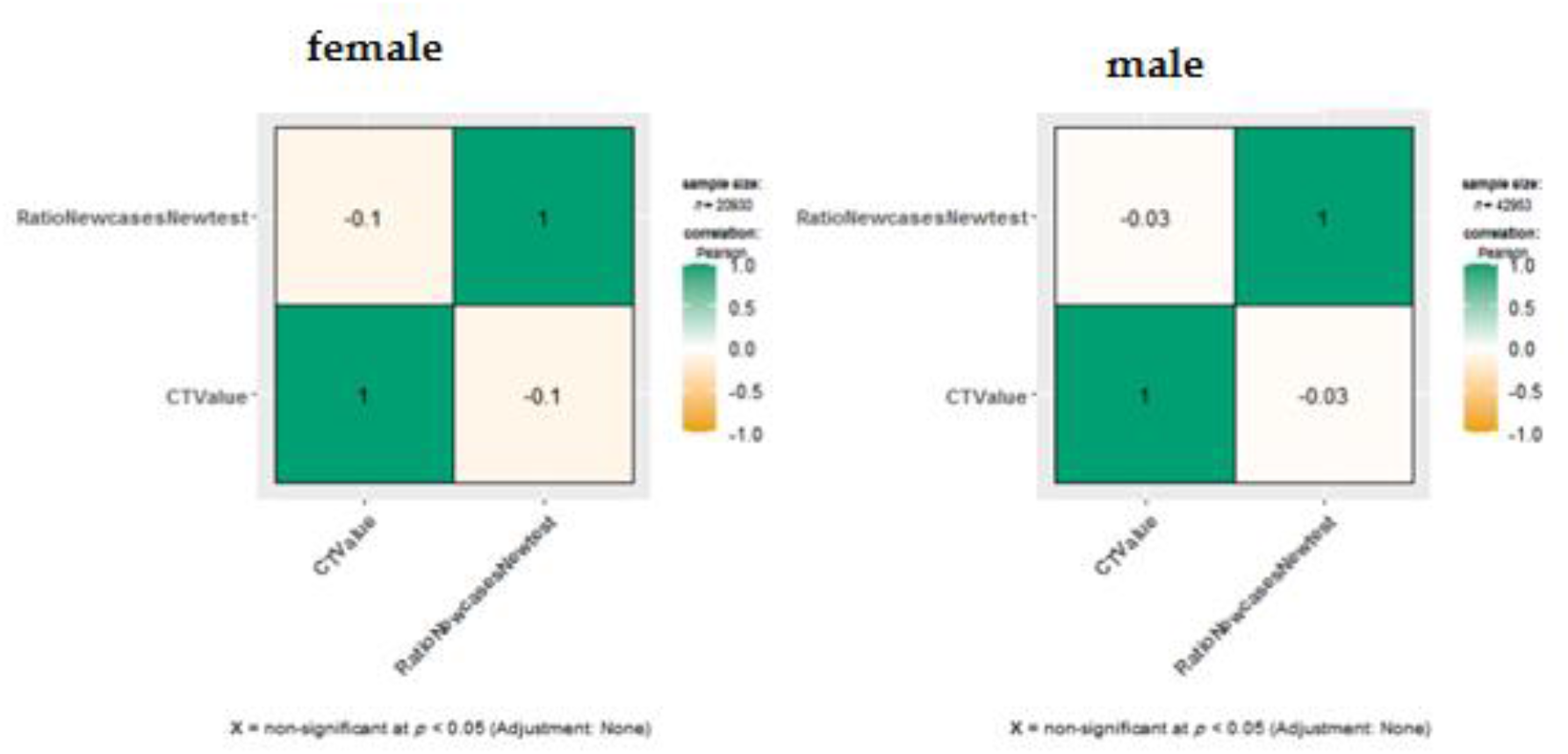
Figure 7 shows the effect of gender on the relationship between Ct values and ratio new cases/new tests. When stratifying for gender, the relationship was found to be negative and significant in females (r = −0.1, p=0.001), but lower and non-significant in men (r = −0.03, p=0.486).

**Figure 8a.**
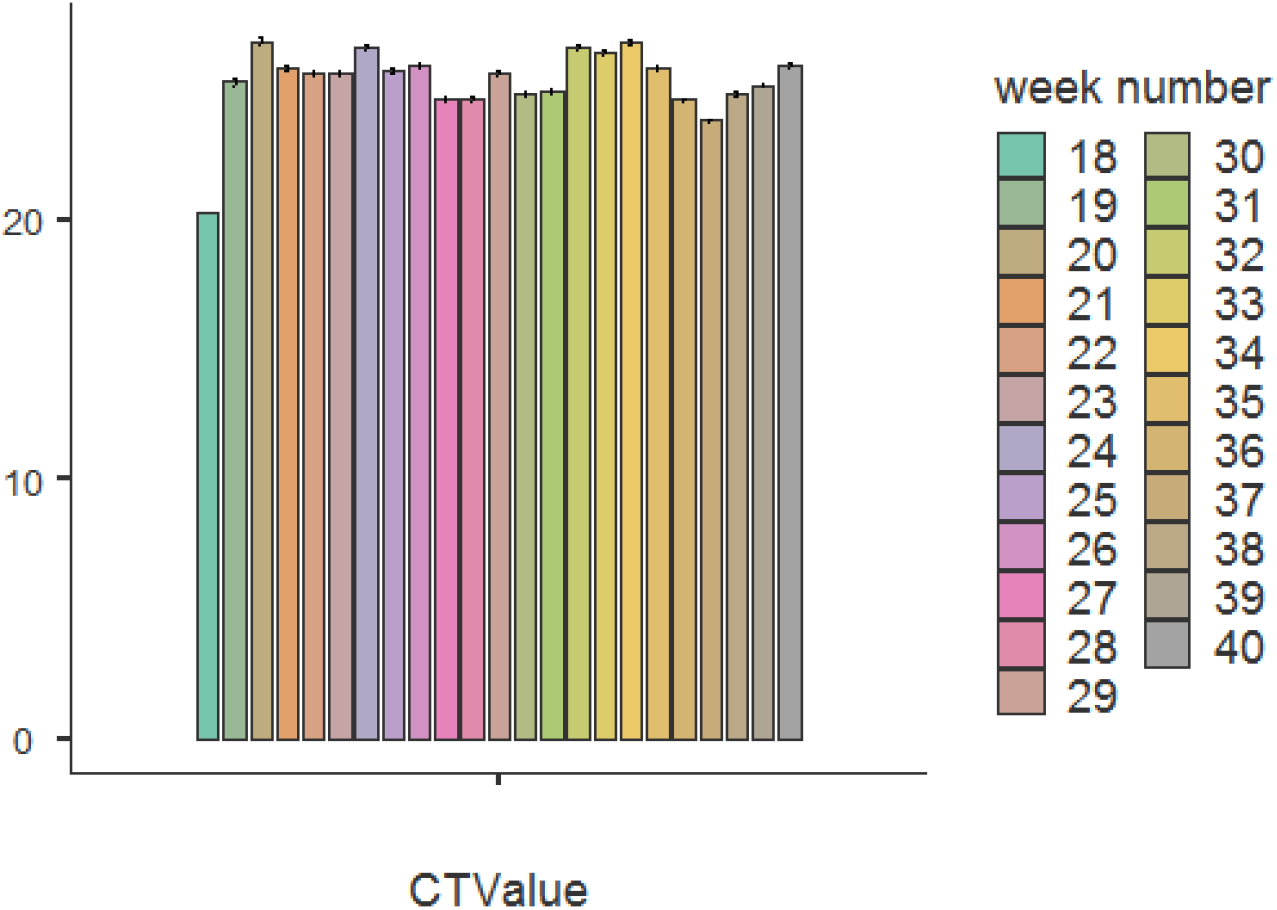
Figure 8a and b show the mean Ct values from week 18 to week 40.

**Figure 8b.**
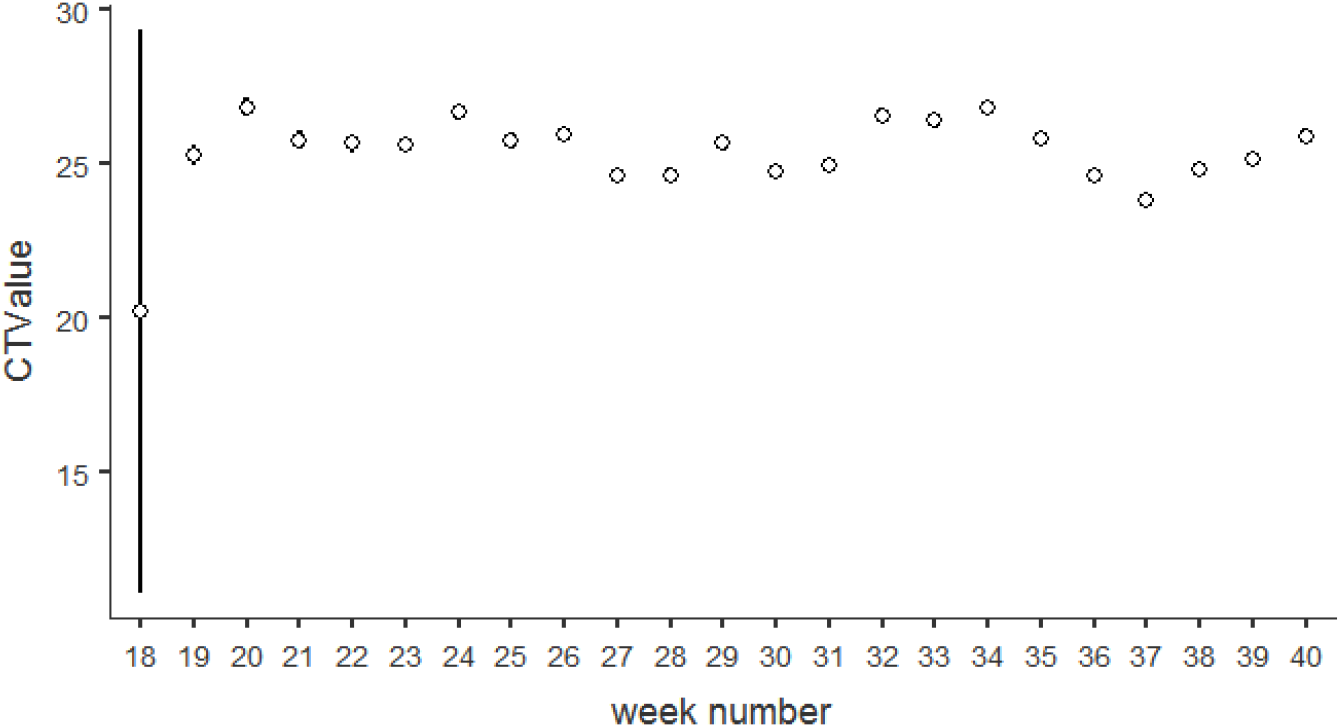
Figure 8b shows the effect of Ct values (adjusted for % of new cases) during the time period (from week 18 to 40). There are no changes regarding the relationship *Ct value and week number (adjusted for % new cases).

**Table 2.** Regression model on the association between Ct values and New daily cases. Model adjusted for regressors: age, new daily tests. The level of prediction of this model is 45.3%, while the Pearson’s correlation coefficient is considered good r=0.673. In this regression model, the age is not associated with the new daily cases, while the associated factors were the Ct values and the number of daily tests. The linear regression equation for the prediction of the new daily cases is: y = (new daily cases) = 65.88+ Ct value *(−1.81)+ daily tests*(0.05).

| **95% Confidence Interval** |  |  |  |  |
| --- | --- | --- | --- | --- |
| **Predictor** | **Estimate** | **Lower** | **Upper** | **p** |
| Intercept | 65.88393 | 59.47637 | 72.29150 | 0.000001 |
| CT Value | -1.81932 | -1.98857 | -1.65007 | 0.000001 |
| Daily Tests | 0.05160 | 0.05117 | 0.05202 | 0.000001 |
| Age | 0.02363 | -0.02407 | 0.07133 | 0.33153 |

### Development of the Predictive Model

Based on the linear regression analysis, the equation for the prediction of the new daily cases is:

New Daily Cases = 65.88+ (−1.81 x Mean Ct Value) + (0.05 x Mean Daily Tests)

When analyzing the relationship between Ct values and ratio of positive tests while stratifying for nationality, the significance of the correlation varies. A negative correlation was found between Bahraini cases (p = 0.001) only. Additionally, when stratifying for gender, the relationship was found to be negative and significant in females (r = -0.1, p=0.001), but lower and non-significant in men (r = -0.03, p = 0.459).

## Discussion

The correlation between RT-PCR Ct values in an infected sample of the population and a change in the overall number of positive cases over time, has not been reported in the literature to date. In this study, Ct values were used as an indicator of viral load and transmissibility. Considering that positive cases with low Ct values have a higher viral load than cases with high Ct values, and that a higher viral load leads to an increase in viral shedding and thus contagiousness in a population, it is reasonable to expect that a decrease in population Ct values would correlate with a surge in cases.

To begin with, our study reported a male to female sex ratio of 2.05:1. This ratio is much higher than the ratio of 1.03:1, reported by the WHO’s global case-based surveillance system^10^. This is mainly due the fact that the 47% of infected cases during the studied period were Non-Bahraini, with majority of whom belong to labor camps, which have a male majority. Moreover, this may also be driven by cultural norms, with males more likely to socially mix outdoors and thus be infected. This may also be related to the higher percentage of males in the population (62.6%), with a male to female ratio of 1.67:1^11^.

Of the cohort, only 35.25% were symptomatic, with 64.75% presenting asymptomatically, one of the highest asymptomatic rates reported in literature to date^12^. The symptom status for these cases was documented upon testing and upon triage; hence a subset of the asymptomatic cases might be further classified as presymptomatic. The prevalence of asymptomatic patients is very difficult to assert, a meta-analysis of 50,155 cases from 41 studies suggests that it may account for 15.6% (95% CI, 10.1%-23.0%) of SARS-CoV-2 infections^13^. The prevalence reported in our cohort may be higher for a number of reasons, including more open screening policies and comprehensive contact tracing. It may also be associated with other factors inherent to cohort characteristics, such as age (which is on the low-end of reported means^14^) and comorbidities. It is worth noting that our study found a correlation between lower Ct values and symptomatic presentation, however the mean Ct value difference between symptomatic and asymptomatic was 0.86 which may not be a clinically significant difference. It may be suggested that a higher asymptomatic prevalence could be indicative of more well-controlled isolation measures leading to a lower viral transmission in the population. It must be noted however that since this is a cross-sectional study of a specific point in time, that some of the cases may be pre-symptomatic rather asymptomatic. Regardless, the prevalence would still be higher than that reported in other cohorts^15^.

According to our findings, as hypothesized, RT-PCR Ct values seemed to be negatively correlated with the number of daily cases. In other words, the lower the Ct value, the higher the fraction of new positive tests in the population. Similarly, a recent population-based study (pre-print) from Massachusetts, USA, has identified a relationship between the population-level cross-sectional distribution of Ct values and the growth rate of the epidemic. The data was used to successfully develop an accurate inference model of the outbreak’s trajectory^16^. A surveillance study from Italy has also reported a significant increase in Ct values over three different consecutive epidemic periods, indicating a possible association with underlying epidemiologic dynamics^17^. Considering evidence showing that every 1-unit increase in Ct values demonstrates a 0.64 odds ratio for positive viral culture (95% CI; 0.49– 0.84; P < .001)^18^, it would be reasonable to infer that the association of lower Ct with a surge in cases is driven by increased viral infectivity in the population. In support of this hypothesis, one study from Singapore found that surface contamination from admitted patients declined with increasing Ct values and duration of illness. No correlation however was found between Ct values of clinical samples and Ct values of environmental samples across days of illness^19^.

Upon stratification by demographic factors, the association between lower Ct values and increase in fraction of positive tests remains the case for all ages, females, and Bahraini-nationality, but does not apply to Males and Non-Bahraini nationalities in our cohort. Possible justifications for these observations are multi-factorial. First, regarding Ct values not being correlated with proportion of positive tests in non-Bahrainis, this may be attributed to the unique characteristics and confounders of this sub-group and the associated targeted public health measures. The large majority of non-Bahrainis in our cohort are composed of migrant workers. A number of observations were made early on in the outbreak that made this sub-group stand out, keeping in mind the often-characteristic background of a lower socioeconomic status. Notably, it was observed that the symptom threshold at which this sub-group would present to the healthcare services was much higher than the Bahraini subgroup. Furthermore, living areas of migrant workers where targeted for COVID-19 screening, often finding whole buildings of positive cases; this may have also been a major contributing factor to the statistically significant higher prevalence of low Ct values in this subgroup. Keeping both of these unique observations in mind, the identification of a correlation via comparison of changes in Ct values and number of cases may have been diluted by the skewed homogenous nature of this subgroup. Furthermore, this subgroup was exposed to unique and intensive public health campaigns that attempted to reduce the household cluttering of migrant workers, and provide health education and awareness. This was largely effective, approaching a standardized state between Bahraini and non-Bahraini subgroups towards the end of the study period. As such, it is clear from our observations that the non-Bahraini subgroup presents with unique, multifactorial confounders and potential factors that may have affected and differentiated the association between Ct value and number of cases. In fact, biologic and genetic/epigenetic factors may have also been at play.

As for the finding regarding the male gender not being associated with Ct value and number of cases, this may be attributed to three main possibilities. The first, and most likely, is that the large majority of non-Bahraini migrant workers were male, which bring us back to the confounders associated with this subgroup. The second possibility is related to the social behaviors of males, who are more likely to mix outside and contract the virus, and less likely to work from home; this may also provide an explanation to the higher prevalence of low Ct values in this subgroup, although not statistically significant. Differences in health behaviors between males and females may also have played a role. Finally, the third possibility would be related to a biologic difference between both sexes. The mechanism of such factors and how they may be related, if at all, remains unclear. More studies in different cohorts while accounting for confounders may help develop a fuller picture.

It is also important to consider that the mean Ct value for the cohort was 25.49 (95%CI: 25.45 -25.52). As such, the Ct values remained largely stable and within a very tight confidence interval, with no statistically significant weekly changes over the time period. The lack of major changes may have not allowed us to view all associations between Ct values and positive cases to the extent to which they may be intertwined in reality, which may have surfaced at a specific threshold or degree of difference. On the other hand, it is also important to consider that the time between testing and acquiring of infection varies from every individual. Therefore, a patient may have acquired a COVID-19 infection during a period of low Ct values in the population clusters, but was only tested positive a while after, when the corresponding Ct values had increased.

Although lower Ct values indicate a higher viral load and thus increased infectiousness, which would be expected to be associated with a surge in cases as was shown in our data, it is important to consider the confounding nature of symptomatic status. For one, asymptomatic status can lead to increased transmission compared to symptomatic, because symptomatic individuals are able to identify themselves self-isolate and get tested as soon as possible. Since asymptomatic cases also had statistically significant higher Ct values according to our findings, it is important to consider that the association between Ct values and infectivity on a population scale may be even more significant than reported. Furthermore, all COVID-19 cases had their Ct values reported to the COVID-19 taskforce. Therefore, those with lower Ct values might have had more aggressive tracing and management, which can indirectly contribute to lowering infectivity from that individual.

Regarding asymptomatic cases presenting with higher Ct values, this may be attributed to a pathophysiologic mechanism whereas a higher Ct value, and thus lower viral load, may decrease or dampen the symptomatic presentation of said infected patient. It is important to also consider that asymptomatic individuals may have had higher Ct values due to being identified at a later time since infection compared to symptomatic individuals. It has been reported that Ct values increases over illness duration^20^.

Change in COVID-19 cases is largely multifactorial, affected by a number of factors that include testing policies, public health measures, population behavior and compliance, religious/cultural events/holidays, reopening strategies and viral factors^21^. For these reasons, Ct values alone may not be useful in predicting a change in COVID-19 cases, despite our study showcasing an association between these variables. However, they may still be of benefit as part of a multivariable model: Especially one that takes into consideration individual characteristics (e.g. symptomatic status, demographic conditions) as related to individual behavior, and population characteristics (e.g. public health policies) as related to population behavior (e.g. compliance to regulations). Additionally, meticulous contact tracing, testing, and quarantining may have contributed to maintaining the Ct values in the middle, with the majority being between Ct 20 and 30.

Regardless, whether these findings would be clinically relevant is debatable. This reaffirms that disease progression is multifactorial. However, it may be more useful as a metric to use as part of a more diverse tool. Additionally, since healthcare professionals interact with infected patients, extra precautions may be justified in the case of patients with lower Ct values, as they would shed more virus particles and are more likely to be more infectious^22^. Additionally, this may help with deciding which patients are better isolated or placed in negative pressure rooms. The main applications of Ct values however would be best concentrated as epidemiologic/public health and forecasting efforts.

From a public health perspective, knowing that average viral loads may be associated with surge in cases and with a more symptomatic presentation, supports the importance of decreasing viral exposure and the gradual nature of infection. This is in contrast to adopting an “all-or-none” attitude of completely eliminating potential exposure. For example, this would translate to emphasizing that the more socially distanced people are, the lower the chance of acquiring COVID-19, as opposed to 6-feet social distancing being reiterated. This may especially be of importance in public campaigns, reinforcing the importance of maximizing distance and minimizing contact and interaction, rather than only reiterating specific instructions for what should and should not be done to avoid contacting COVID-19.

Despite this being one of the largest cohorts on COVID-19 in the region, and the largest to investigate Ct values in relation to case trajectory, the limitations are important to be addressed. To begin with, although this is a multi-center analysis, it remains a national one in a small country. National public health measures and screening policies differ from one place to the other, which necessitates the need for a global cohort when investigating the macro-behaviors and characteristics of COVID-19. The Ct values as reported in our study have been fairly stable throughout the five-month period from a statistical standpoint, which would make it non-ideal to test for associations between Ct values and case trajectory. For that reason, longer periods or cohorts from different populations may be more suitable to investigate an association. Screening policies additionally play a vital role in determining the cohort characteristics. Our study involved cases from varying screening approaches, including admitted patients, symptomatic suspected cases in outpatient setting, contact tracing, population screening in case of travel, random testing in the community and individual-requested testing. In order to minimize bias and confounding factors, ideally only cohorts from randomized population screening should be included. This would also help control the number of tests conducted per day. The different frequencies of testing may have impacted the proportion of positive cases identified per day, contributing to the change in cases in a way that conflicts with Ct values as the intended independent variable. Additionally, public health measures changed reactively throughout the study period, which likely impacted the transmission of the virus in the population in a way that would be difficult to account for.

Furthermore, human bias in tracking down the cases with lower Ct values may have skewed the findings. Additionally, The Ct values can vary depending on how the sample has been collected. For instance, a poorly collected nasopharyngeal swab can have very high Ct values, thus leading to biased findings regarding infectiousness or disease severity. Ct values are also determined by the technical competence of the person performing the test, calibration of equipment, and analytical skills of the interpreters, all of which could have impacted the results. Furthermore, inconsistency in Ct values can result from irregular amplification efficiency which has been observed even when using the same commercial kit; hence, it is recommended to locally determine the limit of detection (LoD) of our tests. To add to that, double delta Ct values were not used in our study, which would have been a more standardized method to report viral loads across different PCR kits and machines. The retrospective observational study design is also in itself an important limitation. Time from symptom onset is also an important variable that could be a confounding factor, however the data on it was difficult to collect. Other confounding variables influencing both Ct values and newly identified cases were difficult to completely account for in the analysis.

## Conclusion

The spread of a viral disease of the like of COVID-19 in a population is multifactorial. Public health measures, testing protocols, population compliance, and viral factors all play a role in the degree of transmission, rate of positive cases, and epidemiologic trajectory. Understanding the correlations of a simple, commonly available measure like Ct values with epidemiologic variables and patient demographics may pave the road for the development of tools to forecast a pandemic’s behavior. Our first-of-its-kind and largest-to-date retrospective multicenter national study points towards a possible correlation between Ct values and the ratio of positive tests in a population. Additionally, symptomatic status was associated with a lower Ct value. This reaffirms the need to develop multivariable models to forecast viral pandemics, possibly including Ct values. Similar studies of a large magnitude in different countries may be able to shed more light on this top

## Data Availability

Available on reasonable request to corresponding author

## Declarations

### Conflict of interest

The authors have declared that no conflict of interest exists.

### Ethics approval and consent to participate

The study was approved by the National COVID-19 Research and Ethics Committee.

### Consent for publication

All authors gave their consent for publication.

### Availability of data and materials

All the data for this study will be made available upon reasonable request to the corresponding author.

### Funding

No funding was received to perform this study.

### Author contributions

AKA, AIA collected the data. AKA, MQ, EJ contributed in the design of the study. SP performed the statistical analysis. SM scripted the first draft. All author contributed to writing the manuscript. All authors have read and approved the final manuscript.

## Notes

### Competing Interest Statement

The authors have declared no competing interest.

